# Young onset diabetes in Asian Indians is associated with lower measured and genetically determined beta-cell function: an INSPIRED study

**DOI:** 10.1101/2021.09.07.21263208

**Authors:** Moneeza K Siddiqui, RM Anjana, Adem Y Dawed, Cyrielle Martoeau, Sundararajan Srinivasan, Jebarani Saravanan, Sathish K Madanagopal, Abirami Veluchamy, Rajendra Pradeepa, Naveed Sattar, Radha Venkatesan, Colin N A Palmer, Ewan R Pearson, Viswanathan Mohan

**Author notes:** Corresponding Authors Dr Viswanathan Mohan, Dr. Mohan’s Diabetes Specialities Centre & Madras Diabetes Research Foundation, No. 6, Conram Smith Road, Gopalapuram, Chennai – 600 086, India, Professor Ewan R Pearson, Division of Population Health & Genomics, School of Medicine, University of Dundee Dundee, DD1 9SY, United Kingdom. These authors contributed equally (Moneeza K Siddiqui, RM Anjana, Ewan R Pearson, Viswanathan Mohan).

## Abstract

**Background:** South Asians have higher risk of type 2 diabetes compared to white Europeans and a younger age of onset. Reasons for the younger age of onset in relation to beta-cell function and insulin sensitivity are under-explored.

**Methods:** Two cohorts of Asian Indians, ICMR-INDIAB (Indian Council of Medical Research-INdia DIABetes Study) and DMDSC (Dr. Mohan’s Diabetes Specialties Centre) and one of white Europeans, ESDC (East Scotland Diabetes Cohort) were used. We examined the comparative prevalence of healthy, overweight, and obese BMI in young onset diabetes. We explored the role of clinically measured beta-cell function in diabetes onset in Asian Indians. Finally, the comparative distribution of a partitioned polygenic score (pPS) for risk of diabetes due to poor beta cell function was examined.

**Results:** Prevalence of young onset with normal BMI was 9.3% amongst white Europeans and 24%-39% amongst Asian Indians. In young diagnosed Asian Indians, after adjustment for family history of T2DM, sex, insulin sensitivity and HDL-c, stimulated C-peptide was 492pmol/mL (IQR: 353,616,P<0.0001) lower in lean compared to obese individuals. Asian Indians have lower genetically determined beta-cell function than white Europeans(P <0.0001). The pPS was associated with age of diagnosis in Asian Indians but not in white Europeans. The pPS explained 2% of variation in clinically measured beta cell function and 1.2%, 0.97%, and 0.36% of variance in age of diabetes amongst Asian Indians with normal, overweight, and obese BMI respectively.

**Conclusions:** Asian Indians have over two times the prevalence of lean BMI in young onset diabetes compared to white Europeans. This phenotype of lean, young onset diabetes appears driven in part by lower beta cell function. We demonstrate that Asian Indians with diabetes also have lower genetically determined beta cell function.

## Introduction

Diabetes prevalence is increasing, with a major burden of morbidity in South Asia. The International Diabetes Federation (IDF) projects that by 2045 there will be 151.4 million indigenous South Asians with type 2 diabetes, almost doubling the present prevalence (1). A recent large-scale population survey, the Indian Council for Medical Research-India Diabetes (ICMR-INDIAB) study, reported the average prevalence of diabetes in India was 7.3% with substantial variation across states(2). This is higher than the current prevalence in Scotland (6.3%), a representative White European population, or most of Europe per IDF estimates. Type 2 diabetes in South Asians is a growing global problem as they also form a substantial part of the diaspora living in western countries. Recent estimates from the United States of America and United Kingdom suggest that South Asians form 1.6% and 6% respectively of the total population (3,4). South Asians living in high income countries are reported to be diagnosed with type 2 diabetes between 5 and 10 years earlier than White Europeans (5–8). To reduce the global burden of diabetes there is a need to better characterise the specific metabolic and physiological drivers of diabetes in this ethnic group.

The metabolic phenotype of the Asian Indian differs markedly from the phenotype seen in whites. Yajnik *et al*. coined the phrase “thin-fat” Indian, best captured in the comparative DEXA (Dual Energy X-ray Absorptiometry) scan of Yajnik and Yudkin (9). This phenotype is of increased truncal fat despite normal or low body mass index (BMI) which is associated with hyperinsulinemia and other measures of insulin resistance, and is seen from birth(10–13), in children(14,15) and through adult life. A number of studies have investigated the association of this thin-fat phenotype with diabetes risk(16–19). Large-scale studies so far have focussed on migrant South Asian populations living in high income countries. Of these, two more recent comparative studies across ethnicities are of note. The SABRE study(19) was a prospective cohort study undertaken in the UK comparing risk for type 2 diabetes and parameters associated with diabetes risk in white, Asian Indian and Afro-Caribbean ethnic groups living in the UK. This study reported that, compared to White Europeans, Asian Indian men who go on to develop diabetes over a 20-year follow-up have lower BMI, higher waist-hip ratio, higher truncal skinfolds, higher insulin resistance, and increased (compensatory) beta-cell function, but with no difference in lipids. More recently, an analysis of UK Biobank established that, to have the same diabetes risk as White participants with a BMI>30kg/m^2^, the equivalent BMI in South Asian participants was only 22kg/m^2^ (20).

While it has been established that Asian Indian type 2 diabetes is characterised by younger age of onset with a relatively low or lower BMI (21–23), there have been no large-scale studies undertaken to comprehensively characterise the metabolic phenotype and genetic risk of young, lean Asian Indians who develop type 2 diabetes. A major challenge in comparative studies of trans-ethnic diabetes is the lack of genetic resources which can be used to overcome biases introduced when comparing clinical measures from different geographic health settings. The India-Scotland Partnership for Precision Medicine in Diabetes (INSPIRED) brings together data resources for the study of diabetes in India and Scotland, UK. We performed the first population study to comprehensively understand the metabolic risk factors for young onset diabetes in an Asian Indian cohort using ethnicity-specific thresholds for obesity. We then test the hypothesis that the phenotype of young onset diabetes in Asian Indians is one of beta cell deficiency using both clinical data and a partitioned polygenic score for type 2 diabetes susceptibility due to impaired beta cell function.

## Methods

### Cohorts used for the INSPIRED study

#### 1. INDIAB Cohort

The Asian Indian cohort is derived from published data of the ICMR-INDIAB (Indian Council of Medical Research-India Diabetes) study (2). This is a national cross-sectional study to establish the national and state-specific prevalence of diabetes and prediabetes in India. Anthropometric and clinical data was surveyed from representative participants belonging to 15 states in India. The survey methodology has been described previously (24). Individuals underwent an oral glucose tolerance test (OGTT) (which included a fasting and 2hr post 75gm load glucose measurement) (25) and were classified as having newly diagnosed diabetes as per the WHO criteria.

#### 2. East Scotland Diabetes Cohort (ESDC)

The White European cohort includes patients from the East of Scotland from population-level data on individuals with type 2 diabetes across the regions of Tayside & Fife in Scotland. Clinical data is made available through the Scottish Care Information-Diabetes Collaboration (SCI-DC) system (26). Type 2 diabetes is diagnosed by general practitioners by the use of fasting or random glucose, oral glucose tolerance test or HbA1c and using WHO criteria for diagnosis (25) and recorded in the SCI-Diabetes system. Newly diagnosed diabetes was defined as individuals with a recorded diagnosis of type 2 diabetes with all anthropometric and clinical measures reported within 1 year of diagnosis.

#### 3. India Clinical Diabetes Cohort-DMDSC

In order to investigate the physiological, biochemical and genetic factors associated with young onset diabetes in Asian Indians we utilised data from Dr. Mohan’s Diabetes Specialities Centre (DMDSC). This is a large, privately-run chain of single-speciality hospitals and clinics for the treatment of diabetes and comorbidities and is a key site for diabetes research in India (2,27,28). DMDSC is a network of 50 clinics, providing data from over 500,000 patients with type 2 diabetes. Data linkage was carried out using a unified electronic health system across clinic, biochemistry, immunology, radiology, retinal screening and other sub-specialties.

### Study Inclusion criteria for DMDSC cohort

The main inclusion criteria for the study was the diagnosis of type 2 diabetes using the WHO criteria for diagnosis (25). All patients in DMDSC cohort had type 2 diabetes and were excluded if clinically diagnosed with type 1 diabetes or if GADA positive during the course of treatment and follow-up at DMDSC. The data used in this study is cross-sectional, however diabetes classifications were applied retrospectively to ensure the population under study had type 2 diabetes. The study period was either 3 months before, or up to 12 months after, the date of diagnosis of type 2 diabetes. All anthropometric measures and biochemical test results are limited to the study period. If there were multiple measures in that time period, the measure closest to the date of diagnosis was used. We restricted our analyses to individuals diagnosed over the age of 18 years. For further details on cohorts used *see* Supplemental Methods 1.

### Clinical data

Clinical variables were analysed only for the diabetes specialty centre for Asian Indians. Clinical variables included age of diagnosis, sex, HbA1c, BMI, systolic and diastolic blood pressure (SBP & DBP), total cholesterol, HDL-cholesterol (HDL-C), LDL-cholesterol (LDL-C), triglycerides, creatinine and liver enzymes. These were measured in the fasted state in. LDL-Cholesterol was calculated using the Friedewald formula.

An advantage of the use of Asian Indian DMDSC cohort included the availability of number of important measures (such as family history of diabetes in first degree relatives, fasting and post-prandial C-peptide, HOMA-B and HOMA-S). HOMA-B and HOMA-S were calculated from fasting C-peptide and glucose using the HOMA calculator (29–31).

### Genetic data

Genetic data from the white European ESDC cohort was available from four platforms: Affymetrix 6.0 Affymetrix, Santa Clara), Illumina OmniExpress -12VI platform and Illumina Infinium (Illumina, San Diego) for a total of 6,933 white Europeans with diabetes (32). Genetic data for DMDSC was available from the Illumina Global Screening Array for 5,806 Asian Indians with type 2 diabetes. Imputation was performed against the Haplotype Reference Consortium panel for both populations (33) using the Michigan Imputation Server and Sanger Imputation Server. Genotype quality threshold of 90% was applied for imputed SNPs. All genotyped individuals had a confirmed diagnosis of type 2 diabetes per criteria described above.

Partitioned or process-specific polygenic scores (pPS) for type 2 diabetes risk were developed by Mahajan *et al*. (34,35). Of the 6 partitioned scores developed, we focus on the beta-cell dysfunction cluster consisting of 8 loci classified as “Insulin secretion 1” which was aetiologically distinct from other causal clusters for type 2 diabetes. The empirical distribution of the unweighted pPS was examined. A greater number of risk alleles implies increased risk for type 2 diabetes due to poor beta cell function. Further, in order to compare the genetically determined differences in beta-cell function based on this pPS, the variants were weighted for their association with surrogate estimate of beta-cell function (HOMA-B) (35). A lower weighted pPS implies lower genetically determined beta cell function. The comparative distribution of the weighted insulin-secretion pPS was examined across the two ethnicities. A sub-group of Asian Indians who had been genotyped and also met clinical study inclusion criteria of diagnosis within 12 months of clinic visit (n=1513) were used to examine the relationship between the insulin secretion pPS, age at diagnosis and BMI.

Replication in the UKBiobank was undertaken using individuals diagnosed with type 2 diabetes (36). Ethnicity was classified using self-reported ethnicity cross-validated with principal components analyses (37). We compared the distribution of the same insulin-secretion pPS risk variants in white British and South Asians.

### Study design

Cross-sectional analyses of data were undertaken using the time point of diabetes diagnosis in all cohorts. Young onset diabetes was defined as diagnosis at or under the age of 40 years (38). Additionally, sensitivity analyses using population-derived age cut-offs were performed. The use of ethnicity-specific BMI cut-offs to define normal, overweight and obese BMI are recommended (39,40). We utilised a consensus definition in Asian Indians recommending thresholds of <23 kg/m2, 23-25kg/m^2^, and >25kg/m^2^ to classify normal, overweight and obese BMI (41,42). Subsequently, a detailed examination of risk factors for young onset diabetes in Asian Indians was undertaken by stratifying on the basis of BMI category in the DMDSC cohort. Finally, a comparison of the pPS for beta cell function in Asian Indians (DMDSC) and white Europeans (ESDC) with type 2 diabetes was undertaken.

### Statistical analyses

The distributions of clinical and anthropometric traits as well as the pPS were assessed for normality. Means and standard deviations were computed for normally distributed variables and medians and inter-quartile ranges were computed for all non-parametric variables. Tests for trend across age and BMI groups was performed. Test for trend for parametric variables were performed using linear regression (T statistic), non-parametric using Jonckheere-Terpstra test (Z statistic) and for categorical variables using Cochran-Armitage Trend test (Z statistic). Quantile regression analyses were undertaken to detect differences in glycemic traits across BMI groups in models adjusted for sex, family history, HDL-cholesterol, and insulin sensitivity. Finally, the comparative distribution of the pPS for beta cell function between Asian Indians and white Europeans with type 2 diabetes was assessed using the Wilcoxon-Mann-Whitney test for differences in median and Kolmogorov-Smirnov statistic for testing two-sample non-parametric distributions. Similarly, a replication in the UKBiobank comparing the distribution of the pPS in white British and South Asians with type 2 diabetes was performed. Jonckheere-Terpstra Z statistics were used to assess in the relationship between the pPS and age groups at diagnosis. Generalised linear models were used to test the association of the pPS with age of diagnosis. Statistical analyses and graphs were produced using SAS 9.4 (SAS Institute, Cary, North Carolina). Values are reported in SI units throughout.

## Results

1,712 Asian Indians from ICMR - INDIAB, 54,989 Asian Indians from DMDSC and 42,563 white Europeans from ESDC all with newly diagnosed type 2 diabetes were categorised by their age at diagnosis and BMI (Figure 1). Characteristics of these three populations are described in Supplementary Table 1. Characteristics of these three populations are described in Supplementary Table 1. The average age of diabetes diagnosis amongst Asian Indians in INDIAB was 50 years and in DMDSC was 47 years, and in white Europeans was 62 years (Supplementary Figure 1).

**Figure 1.**
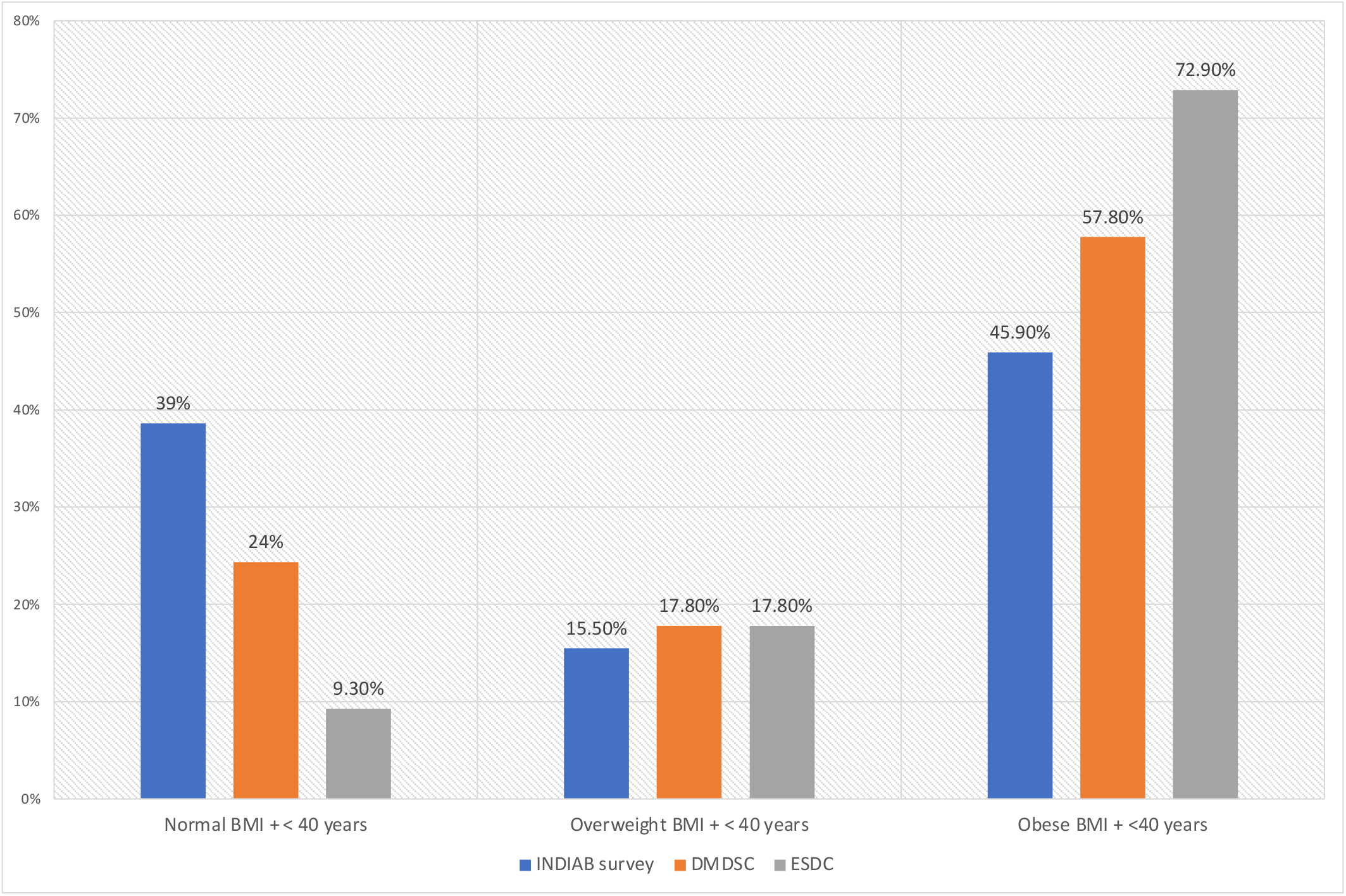
Bar graphs of Asian Indians (blue -INDIAB and orange-DMDSC) and white Europeans (grey-ESDC) with early onset diabetes by proportion belonging to each BMI category. Normal BMI for Asian Indians was <23 kg/m2, overweight: 23-25kg/m2, obese: >25kg/m2. Normal BMI for white European population is <25 kg/m2, overweight 25-30 kg/m2 and obese >30 kg/m2 [16].]. Early onset for both ethnicities was defined as those diagnosed at 40 years or younger (Further data on cohorts available in Supplementary Figure 1 and Table 1)

Early onset diabetes with normal BMI occurs at over twice the frequency in Asian Indians compared to white Europeans

Ethnicity-specific cut offs for normal, overweight and obese BMI were applied. As seen in Figure 1, the majority of Asian Indians diagnosed young, 45.9% in INDIAB and 57.8% in DMDSC had obese BMI (>25kg/m^2^). Similarly, the majority of white Europeans 72.9% had obese BMI (>30 kg/m^2^). However, a shift of distribution was observed in young onset diabetes with normal BMI. 39% of Indians from the INDIAB study and 24% of Indians from DMDSC were classified as having normal BMI (< 23 kg/m^2^), in contrast with 9.3% of white Europeans (<25kg/m^2^) (Chi-sq=25,P<0.0001). A sensitivity analysis using population-derived age cut-offs showed similar results (Supplementary Figure 2) (Chi-sq=9.8, P value=0.001).

Asian Indians diagnosed young have lower beta-cell function compared to those diagnosed older

Individuals from DMDSC were divided into 2 age groups: young diagnosed (individuals with age of diagnosis of 40 years or younger) and older diagnosed (diagnosis over the age of 40 years). The distribution of clinical and anthropometric features across age groups is described in Table 1.

**Table 1. .** Comparison of characteristics between young and older diagnosed Asian Indians

| **Clinical characteristics** | **Young diagnosed (<=40 years)** | **Older diagnosed (> 40 years)** |
| --- | --- | --- |
| **Numbers** | 15,236 | 39,726 |
| **Age at diagnosis (years) (mean, SD)** | 34.1 (4.35) | 52.6 (8.6) |
| **Sex (% female) \*** | 29% | 41% |
| **Sex (% male)** | 71% | 59% |
| **BMI (kg/m<sup>2</sup>) (mean, SD)** | 26.7 (4.5) | 26.7 (4.4) |
| **Waist circumference (females) (cm) (mean, SD) \*** | 90.4 (10.9) | 91.0 (10.4) |
| **Waist circumference(males) (cm) (mean, SD) \*** | 93.5 (10.6) | 95.2 (10.3) |
| **% males with healthy waist circumference \*** | 54% | 49% |
| **% females with healthy waist circumference \*** | 39% | 35% |
| **HbA1c (%) (mean, SD) \*** | 9.11 (2.3) | 8.7 (2.3) |
| **HbA1c (mmol/mol) (mean, SD) \*** | 76.08 (25.6) | 71.8 (25.0) |
| **Fasted c-peptide (pmol/mL) (median, IQR) \*†** | 1000 (600) | 1000 (500) |
| **Stimulated c-peptide (pmol/mL) (median, IQR) \*†** | 2100 (1500) | 2600 (1600) |
| **Stimulated c-peptide adjusted for insulin sensitivity (median, IQR) \*†** | 606 (485) | 730 (490) |
| **HOMA-S (median, IQR) \*†** | 35.4 (20.6) | 35.1 (18.3) |
| **HOMA-B (median, IQR) \*†** | 47.5 (56.9) | 68.5 (71.4) |
| **Family history of diabetes (%) \*** | 69% | 44% |
| **LDL-c (mmol/L) (mean, SD) \*** | 2.98 (0.91) | 3.04 (0.93) |
| **HDL-c (mmol/L) (mean, SD) \*** | 0.995 (0.22) | 1.06 (0.25) |
| **Triglycerides (mmol/L) (median, IQR) \*†** | 1.76 (1.33) | 1.63 (1.04) |
| **Total Cholesterol (mmol/L) (mean, SD)** | 4.95 (1.14) | 4.98 (1.13) |
| **ALT (IU/L) (median, IQR) \*†** | 32 (27) | 24 (17) |

*\*differences were statistically significant (P<0.05) † statistical test performed on log transformed variable Waist circumference guidelines for Asian Indian males 90 cm and for females 80 cm.*

Both fasted and stimulated C-peptide were significantly lower in those diagnosed young compared to older. Beta-cell function determined using HOMA-B, along with fasted and stimulated C-peptides were also lower in young compared to older diagnosed individuals. A family history of diabetes was observed more frequently in younger individuals. Amongst those diagnosed young a higher proportion were male (71%) compared to female (29%), in spite of more males having normal/healthy waist circumferences (54%) than females (39%). No difference in average BMI was observed between the two groups. Comparative dyslipidemia characterised by higher triglycerides and lower HDL-C were observed in young compared to older individuals, however LDL-C levels were lower in young individuals. Liver enzyme, alanine transaminase (ALT) was significantly higher in younger diagnosed individuals. Overall, a pattern of poor beta cell function along with increased adiposity, and adverse lipid and liver profiles was observed in young Asian Indians with diabetes. These features were then examined across BMI categories amongst those diagnosed young (Supplementary Table 2&3).

Asian Indians diagnosed young with normal BMI have lower beta-cell function compared to those diagnosed young with overweight and obese BMI

An examination of trends across BMI categories in these age groups is provided in Supplementary Table 2. In young diagnosed individuals, those with normal/lean BMI had lower fasted and stimulated C-peptides compared to overweight and obese individuals. (Figure 2). This effect persisted when stimulated C-peptides were adjusted for insulin sensitivity (HOMA-S). Similarly, HOMA-B was lower in the lean compared to overweight and obese groups, while inversely HOMA-S was higher (Supplementary Figure 3). Young diagnosed individuals with normal BMI had lower triglycerides and higher HDL-C compared to those diagnosed young and overweight or obese (Supplementary Figure 4).

**Figure 2.**
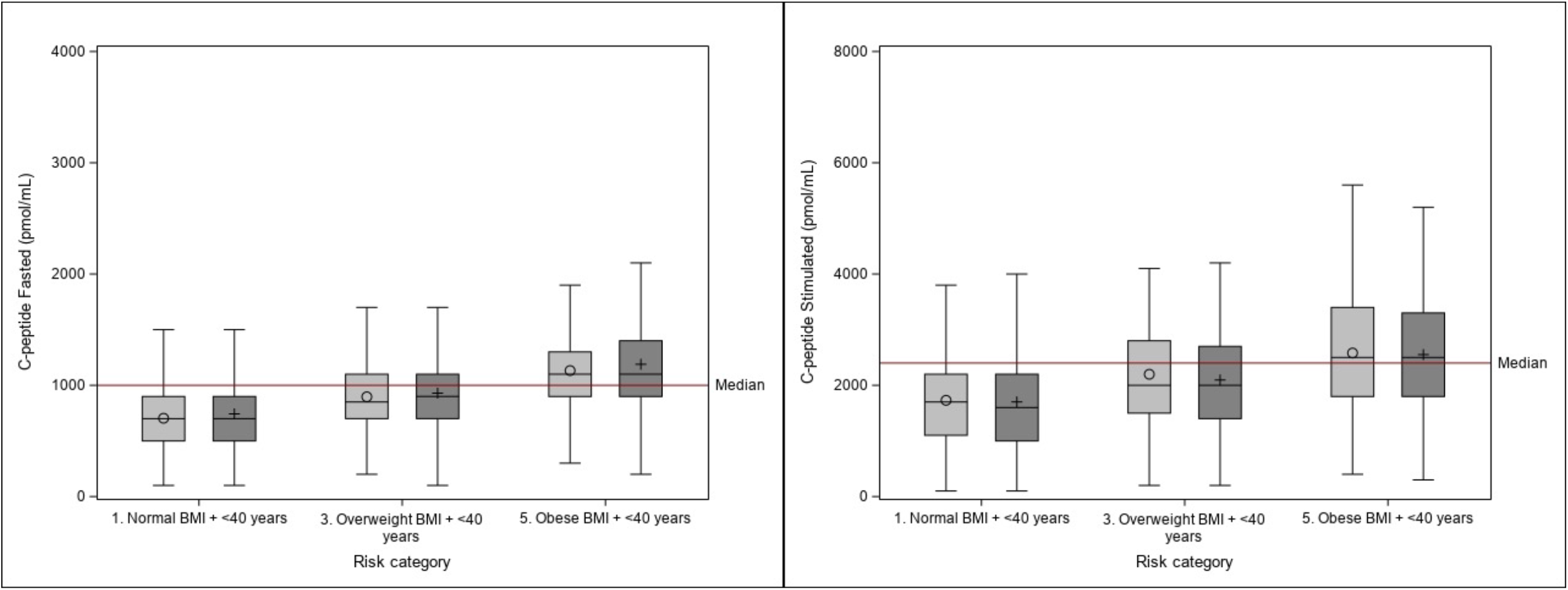
Boxplots demonstrating lower fasted (Figure 2a) and stimulated (Figure 2b) C-peptides by BMI category. Males in light gray, females in dark gray. Those with normal BMI compared to overweight and obese BMI had lower fasted and stimulated C-peptide and similarly lower beta-cell function, with a compensatory increase in insulin sensitivity. When adjusted for insulin sensitivity the association between stimulated C-peptides and age at diagnosis remained significant (P <0.0001).

Together this suggests that diabetes in young lean Asian Indians is not selectively associated with dyslipidemia, but more likely with impaired or poor beta cell function. The result of impaired beta cell function in young lean Asian Indians persisted after adjusting for sex, family history of diabetes, HDL-C, and insulin sensitivity (Supplementary Table 3). The adjusted difference in stimulated C-peptides between those diagnosed with lean compared to obese BMI was 492 pmol/mL (IQR:353,616). The adjusted difference in untransformed HOMA-B was 25.16 (IQR:20.6,28.6).

Partitioned polygenic score for type 2 diabetes risk due to poor beta cell function is lower in Asian Indians compared to white Europeans

We applied the ‘insulin secretion’ pPS of variants in 8 genes associated with type 2 diabetes risk due to poor beta cell function: *MTNR1B, HNF1A, GCK, TCF7L2, TMEM258, ADCY5, SLC3OA8 and ABO* (34,35). Risk alleles across these variants were summed for both ethnicities allowing the comparison of an unweighted partitioned risk score (pPS) (Supplementary Table 4 and Figure 5). There were 5806 Asian Indians and 6933 white Europeans respectively included in this analysis. Compared to white Europeans, Asian Indians had a higher number of risk alleles (Figure 3a, P<0.0001). The difference in the empirical distribution of risk alleles between the two ethnicities was significant (Kolmogrov-Smirnov Pvalue <0.0001). This difference was replicated in UK Biobank using white Europeans (n=17,183) and South Asians (n=1,153) with type 2 diabetes (Supplementary Figure 6a, P<0.0001). The pooled distributions of these risk alleles across the INSPIRED and UK Biobank cohorts is provided in (Supplementary Figure 6b).

**Figure 3.**
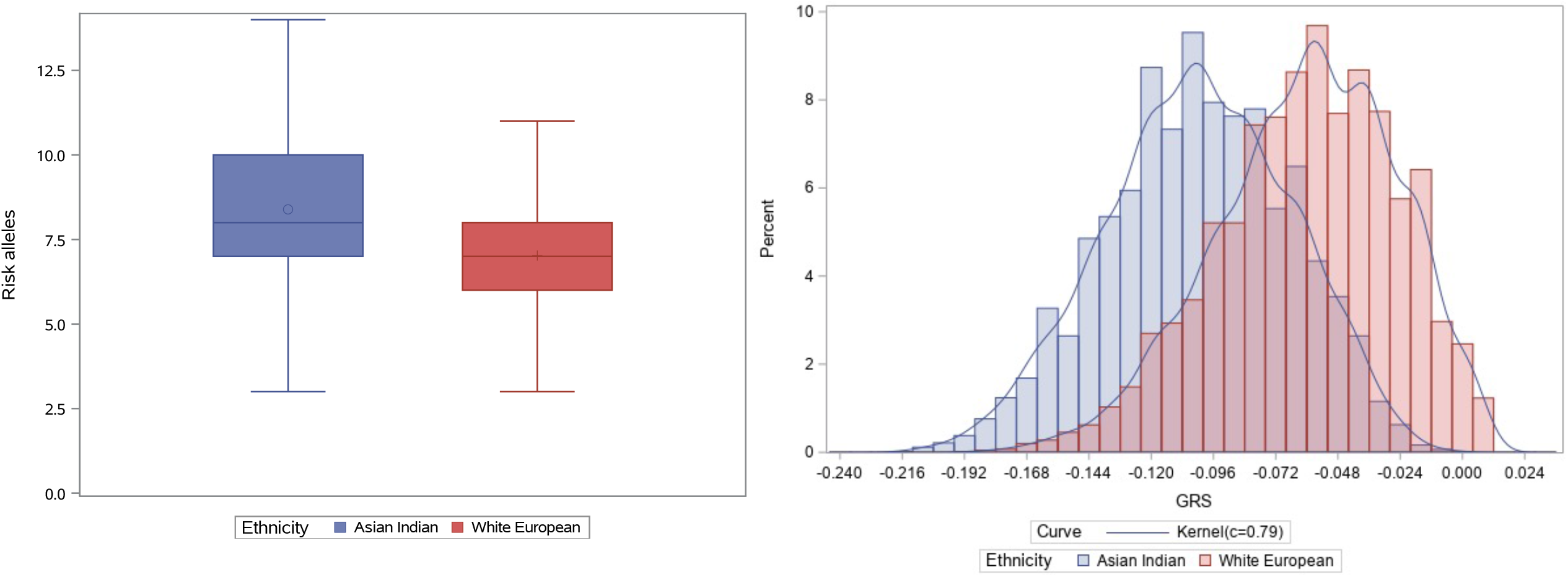
Comparative distribution of beta cell function partitioned polygenic score (pPS) between Asian Indians (in blue) and white Europeans (in red). On the left 3a is a boxplot of the distribution of risk alleles (unweighted pPS) in each group (Wilcoxon-Mann-Whitney P value <0.0001). The empirical distribution of unweighted pPS showed a significant difference using the Kolmogorov-Smirnov asymptotic two-sample test (P value<0.0001). On the right 3b is a histogram showing differences in the distribution of weighted pPS. The difference in distribution was also significantly different (Wilcoxon-Mann-Whitney P value<0.0001).

In the Indian and Scottish cohorts, the risk alleles were then weighted for their association with HOMA-B (Figure 3b) to determine the genetically determined beta-cell function associated with these variants. A lower weighted pPS corresponds to lower HOMA-B and beta cell function. The weighted pPS was found to be non-parametrically distributed in both populations (Supplementary Table 5). A comparison showed Asian Indians with diabetes have lower genetically determined beta cell function compared to white Europeans with diabetes (Figure 3b, P< 0.0001). The weighted pPS was then examined for association with age of diabetes diagnosis (Supplementary Figure 7).

Higher pPS was strongly associated with increasing age of diagnosis in Asian Indians (Z=5.2, P<0.0001), while no trend or association was observed in white Europeans (Z=1.1, P=0.15). We observe that in Asian Indians the trend for the pPS across age groups mirrors that of stimulated C-peptides, HOMA-B and beta-cell function adjusted for insulin sensitivity (Supplementary Figure 8) wherein, those with onset at younger ages have lower stimulated C-peptides, HOMA-B, and genetically determined beta-cell function than individuals with older onset diabetes.

A sub-group of the genotyped Asian Indian cohort who met the clinical study inclusion criteria i.e. for whom BMI was measured at diagnosis of type 2 diabetes was available (n=1513) We used linear regression models stratified by BMI to predict age at diagnosis using the pPS in Asian Indians. We observed that the maximum variance explained was in those with normal BMI (R^2^=1.2), with decreasing variance for overweight (R^2^=0.9%) and obese (R^2^=0.04%) individuals (Supplementary Figure 9 and Table 6).

## Conclusion

### Summary of key results

This study highlights that the age of diabetes onset in Asian Indians is dramatically lower than in White Europeans and that young onset is characterised by a comparatively higher (2.5-4 times) prevalence of individuals with normal BMI using ethnicity-specific thresholds.. Further, we show that amongst Asian Indians, this group (normal BMI + young onset) has lower beta cell function. Using an insulin secretion partitioned type 2 diabetes polygenic score (pPS) we demonstrate that Asian Indians have a higher genetic burden of T2DM risk variants that reduce beta cell function compared to white Europeans. Finally, we demonstrate that the pPS is able to predict age of diabetes diagnosis in those with normal or healthy BMI (R^2^=1.2%) better than in those with overweight or obese BMI. This highlights the role of impaired beta cell function in young onset diabetes in lean Asian Indians.

While this study is focussed on Asian Indians, many studies have used South Asian populations which can include people from Afghanistan, Pakistan, India, Bangladesh, Nepal, Bhutan and Sri Lanka. This is an important distinction as genetic studies have shown a significant heterogeneity in these populations which could be reflected in different clinical phenotypes (43,44). Additionally, thresholds for adiposity are also considered to be different in these ethnic sub-groups (41). This should be considered while evaluating comparisons between our study and others, in addition to the fact that we are examining a non-migrant population.

This is the first study to compare non-migrant populations of white Europeans and Asian Indians and demonstrate the comparative prevalence of lean or normal BMI in those diagnosed young using ethnicity-specific BMI thresholds. We do this by utilising a single threshold of young onset diabetes (diagnosis at age of 40 or younger) and by ethnicity-determined mean age of diagnosis (Figure 1 & Supplementary Figure 2). Our results show using both approaches there exists a preponderance of younger onset diabetes in Asian Indians having normal or healthy BMI. This difference is particularly striking as the average age of diabetes onset is 12 years lower in Asian Indians, demonstrating that diabetes is occurring earlier and in leaner individuals in this ethnic group compared to whites. This is the first time this phenomenon has been clearly documented in non-migrant Indians. It is possible that the lower average age of the underlying Indian compared to the Scottish population could be driving the differences in age at diagnosis. However, considering that these differences have also been documented in migrant Asian Indian and South Asian populations suggests that population structure alone is not driving the observed difference in age at diagnosis.

The phenomenon of lean, young onset is particularly pronounced amongst Asian Indian males, who are more likely than females to have healthy waist circumferences when diagnosed young. There is no clear evidence of dyslipidemia in the young onset normal weight group compared to those with higher BMI. Given the reciprocal relationship between beta-cell function and insulin sensitivity it is not surprising that the insulin sensitivity based on the surrogate estimate of HOMA-S was higher in the lean young group compared to the obese young group. The genetic studies establish that it is the beta-cell deficiency that is causal, with the increased HOMA-S secondary to this.

It is worth noting, that the substantive proportion of young onset diabetes in Indians occurred in those with obese BMI (54% in INDIAB and 57% in DMDSC), this group also have lower insulin sensitivity and higher ALT levels. This is suggestive of higher ectopic fat deposition as ALT is a reasonable correlate of ectopic fat consistent with the hypothesis of “thin-fat” phenotype which has been hypothesized to predispose Asian Indians to type 2 diabetes (9,23,45). This mirrors the more classic insulin resistance aetiology of diabetes.

However, our results suggest that the preponderance of young and lean Asian Indians developing type 2 is driven by poor beta-cell function. The demographics of India are changing – with more urbanisation and a shift to sedentary jobs (46). It is likely that in the background of poor beta-cell function and lower muscle mass, smaller increases in ectopic fat and adiposity could drastically increase diabetes risk. Furthermore, the trend with family history, which is strongly associated with obesity in our data, could be suggestive of a shared genetic risk for obesity, dyslipidemia etc. but could also be reflective of shared lifestyle. Addressing the diabetes epidemic in India and South Asia more broadly, will require efforts in diet-awareness, exercise etc. similarly to those seen in high income countries in addition to correctly diagnosing and treating those developing diabetes due to beta cell impairment.

### Limitations

As yet, there is no conclusive genetic study of beta cell function, HDL-c, obesity, lipodystrophy etc. in individuals of Asian Indian or more broadly, South Asian ancestry. This prevents us from utilising an ethnicity-specific polygenic risk score. It is possible that such a study would produce a different or expanded set of candidate SNPs for beta cell function, since the pPS used in this study has been derived from studies in white European populations. Furthermore, studies on causal genetic variants of beta cell function are not available. The SNPs used have so far not been causally linked to beta cell function, instead they have been shown to be statistically associated with type 2 diabetes risk due to poor beta-cell function (34). In our data, 322 Asian Indians had both c-peptides and genetic data available. We observe that both the unweighted and weighted pPS were correlated with adjusted C-peptide levels (R_unweighted_=12%,P=0.04 and R_weighted_=14%, P=0.01). Notably, these SNPs had no pleiotropic effects on other glycemic traits or diabetes risk factors making them powerful genetic instruments. If these SNPs are not causal variants, they are tagging the causal variants through high linkage disequilibrium (LD). It is possible that the LD structure in these regions could differ between individuals of European and Asian ancestry. However, recently a trans-ancestral study of glycemic traits has replicated many of these signals while reporting that overall, 80% of SNPs identified in single-ancestry studies of glycemic traits showed no evidence of between-ancestry heterogeneity (47). The association of the partitioned polygenic risk score with age of diagnosis in Asian Indians, while mimicking the trend of stimulated C-peptide and HOMA-B strongly indicates its validity in this non-European cohort. In our data, the allele frequencies of the variants show differences across the two ethnicities (Supplementary Table 4 and Figure 4). We observe that out of the 8 variants, Asian Indians had a higher risk allele frequency for 6. This difference in burden of risk alleles was observed in the replication cohort of individuals with type 2 diabetes in the UK Biobank.

Due to the lack of routine screening for diabetes in the Indian population, HbA1c at diagnosis are high. It is likely that the true age of diabetes onset is lower that the age we have recorded. That noted, this differential bias is likely to underestimate our results of early age of diabetes onset with lean BMI.

Another limitation is the lack of comparison of glycemic traits between the two ethnicities particularly C-peptide levels. However, we do not expect to see a different relationship between BMI (as a measure of adiposity) and glycemic traits in white Europeans than we do in Asian Indians i.e. diabetes onset with low BMI is likely to be associated by beta cell failure (48). Our conclusion is that lower beta cell function is observed more frequently in Asian Indians with type 2 diabetes than white Europeans. The comparative genetic study undertaken supports this. Finally, a recurring hypothesis in the aetiology of type 2 diabetes in South Asians involves low lean or muscle mass (21,23). Indeed, studies have found that South Asians have low grip strength compared to white Britons (49). Unfortunately, we do not have the data to examine the relative contribution of low lean mass to young onset diabetes in this study. However, this is a key area of research to be undertaken.

### Strengths

The availability of an intensively phenotyped cohort of non-migrant Asian Indians is novel. The availability of glycemic traits such as fasted and stimulated c-peptides, anthropometric traits such as BMI, waist circumference, lipid traits and family history are unique. The added availability of genetic data alongside these clinical features allows for a definitive exploration of risk factors for young onset disease in this ethnic group. The comparative distribution of the pPS between a population of white Europeans and Asian Indians demonstrates an unbiased estimate of the genetic burden of lower beta-cell function in Asian Indians. Furthermore, the observed difference in burden of risk alleles was replicated using the UK Biobank, where the broader ethnic group of South Asians were compared to white British individuals. The results are consistent with the prevailing hypothesis that beta-cell function is lower in Asian Indians and more broadly in South Asian populations (21,23) which predisposes them to diabetes even at low BMI.

### Generalisability

The data are from a private healthcare centre in India and are more likely to reflect the situation in middle- and upper middle-class residents of urban India. However, we have no evidence that the underlying genetic architecture of urban and rural India are different. Therefore, we expect that the phenotype of diabetes driven by lower beta cell function will persist in rural areas. Indeed, this is clearly evidenced by the high rates of prediabetes prevalence observed in these settings. Moreover, we have also included data from the ICMR-INDIAB Study which is a national survey of diabetes and prediabetes prevalence in a representative population of India and found a nearly equivalent age-adjusted prevalence of prediabetes in urban and rural-dwelling Indians of around 10% (2).

### Interpretation

These results suggest that lower clinically and genetically determined beta cell function contributes to the higher burden of younger onset type 2 diabetes in Asian Indians. In contrast with clustering analyses in white Europeans, the analysis in Asian Indians showed four distinct clusters only two of which overlapped with white Europeans. The Asian Indian-specific clusters were insulin resistant obese diabetes or IROD and combined insulin resistant and deficient diabetes or CIRDD) (50). Similar to the results presented here, this suggests a specific burden in young Indians who develop diabetes due to loss of beta cell function often on a background of obesity. We provide evidence that lower measured and genetically determined beta-cell function is a factor in young onset diabetes in Asian Indians. These findings provide evidence that both the aetiology and presentation of diabetes are somewhat different in Asian Indians than those understood to exist in white Europeans. These results highlight the value of an ethnicity-informed approach to precision medicine in diabetes care.

## Data Availability

The data are available subject to approval by the GoDARTS data access committee and to the Madras Diabetes Research Foundation and can be made by contacting the corresponding authors.

## Ethical committee approvals

India: NIHR Global Health Research Unit on Global Diabetes Outcomes Research, Institutional Ethics Committee of Madras Diabetes Research Foundation, Chennai, India. IRB number IRB00002640. Granted 24^th^ August 2017.

Tayside, Scotland: Ethical approval for the study was provided by the Tayside Medical Ethics Committee (REF:053/04) and the study has been carried out in accordance with the Declaration of Helsinki.

## Author contributions

MKS, ARM, developed the concept and design of the research, performed all analyses, prepared the first draft of the manuscript and revised the manuscript for intellectual content. ERP and VM guided the reviewed and edited the manuscript. AYD, CM, SS, JS, SKM, AV, RP, NS, RV and CNAP contributed to data collection, data access, quality control of data, analysing and interpreting results and revising the manuscript critically for intellectual content. All authors had final approval of the version to be published.

## Data guarantors

Prof. Ewan R Pearson & Dr. V Mohan is the guarantor of this work and as such had full access to all the data in the study and take full responsibility for the integrity of the data and the accuracy of the data analyses.

