## Supplementary materials for "Young onset diabetes in Asian Indians is associated with lower measured and genetically determined beta-cell function: an INSPIRED study"

#### Supplemental Cohort information

#### INSPIRED-East Scotland Diabetes Cohort (ESDC)

In Scotland, all clinical data for patients with diabetes is captured on an electronic medical record system: Scottish Care Information - Diabetes Collaboration (SCI-Diabetes). As all patients with diabetes are managed within the National Health Service, there is complete capture of patient data for all patients from diagnosis. The East of Scotland (Tayside and Fife, population 800,000) has approximately 15 years of complete population capture of prescription encashment. The population included in this study was therefore patients diagnosed with type 2 diabetes after 1994 from Tayside and Fife, who met all other study criteria.

Controls (individuals who did not have diagnosis of type 2 diabetes) were sourced from the GoDARTS study (1) where 8157 controls were recruited between October 2004 and May 2009. Anthropometric lifestyle measurements were recorded for this group at the time of recruitment. Additionally, all patients attend a clinic at recruitment where biochemistry tests are performed, this is the source of the HDL data reported.

All electronic health records were anonymised and made available through the Health Informatics Centre (HIC) in Dundee, with all analysis undertaken on the ISO270001 approved HIC data safe haven.

#### INSPIRED- INDIAB: Asian Indian national cohort

The ICMR-INDIAB study was performed using a multi-stage reporting cross-sectional (2). Data used for this analysis are from the first two phases and the north east phase of this population-survey, performed between November 2008 and July 2015 (3). Capillary oral glucose tolerance tests were used to diagnose diabetes per WHO criteria.

As part of this survey to estimate the prevalence of diabetes and pre-diabetes a variety of anthropometric, clinical, socio-economic and lifestyle factors were recorded. We report findings data from 14 Indian states (out of 28) and 1 union territory.

#### INSPIRED - DMDSC: Asian Indian diabetes-specialty cohort

Dr Mohan’s Diabetes Speciality Centre (DMDSC) is a privately-run chain of single speciality hospitals and clinics for treatment of diabetes and associated disorders. The institution has been in operation since 1991 and currently there are 50 clinics in various locations across 10 states in India, all connected through a single electronic medical system. At present, there are approximately 500,000 patients with Type 2 diabetes within DMDSC, who comprise our Asian Indian cohort; 55,148 whose first clinic visit was within 1 year of diagnosis are presented here.

Patients registered any of the clinics are given a unique ID which is linked to the common Diabetes Electronic Medical Records. All investigations done including biochemical testing, imaging, anthropometric measurements, retinal exams, doppler, biothesiometry or surgical treatments can be viewed by physicians across clinics giving the patient freedom to access any clinic of his/her choice. All test results are directly fed into the system. Physician interaction notes and details of prescriptions issued are available in the system. Pharmacy records are also fed into the system with all the above linked to the unique patient ID. The Madras Diabetes Research Foundation (MDRF) is the research wing of DMDSC and helps in data management and mining. The DMDSC laboratory is accredited by College of American Pathologists (CAP) as well as by the National Accreditation Board for Testing and Calibration Laboratories (NABL) for measurement of glucose, insulin and c-peptide.

#### Clinical variables

HOMA in the DMDSC cohort was derived using fasting c-peptide in patients who were diet treated, treated with oral antihyperglycaemics, and in a small percentage, with insulin. Comparison was made with white European participants from the ADOPT trial (4) and the UKPDS trial (5,6). Since different assays have been used to measure fasting glucose and insulin or c-peptide in these populations the HOMA results have been plotted separately to highlight the difference in overall trend, not the absolute difference in levels (Supplementary Figure 6).

### Supplementary Results

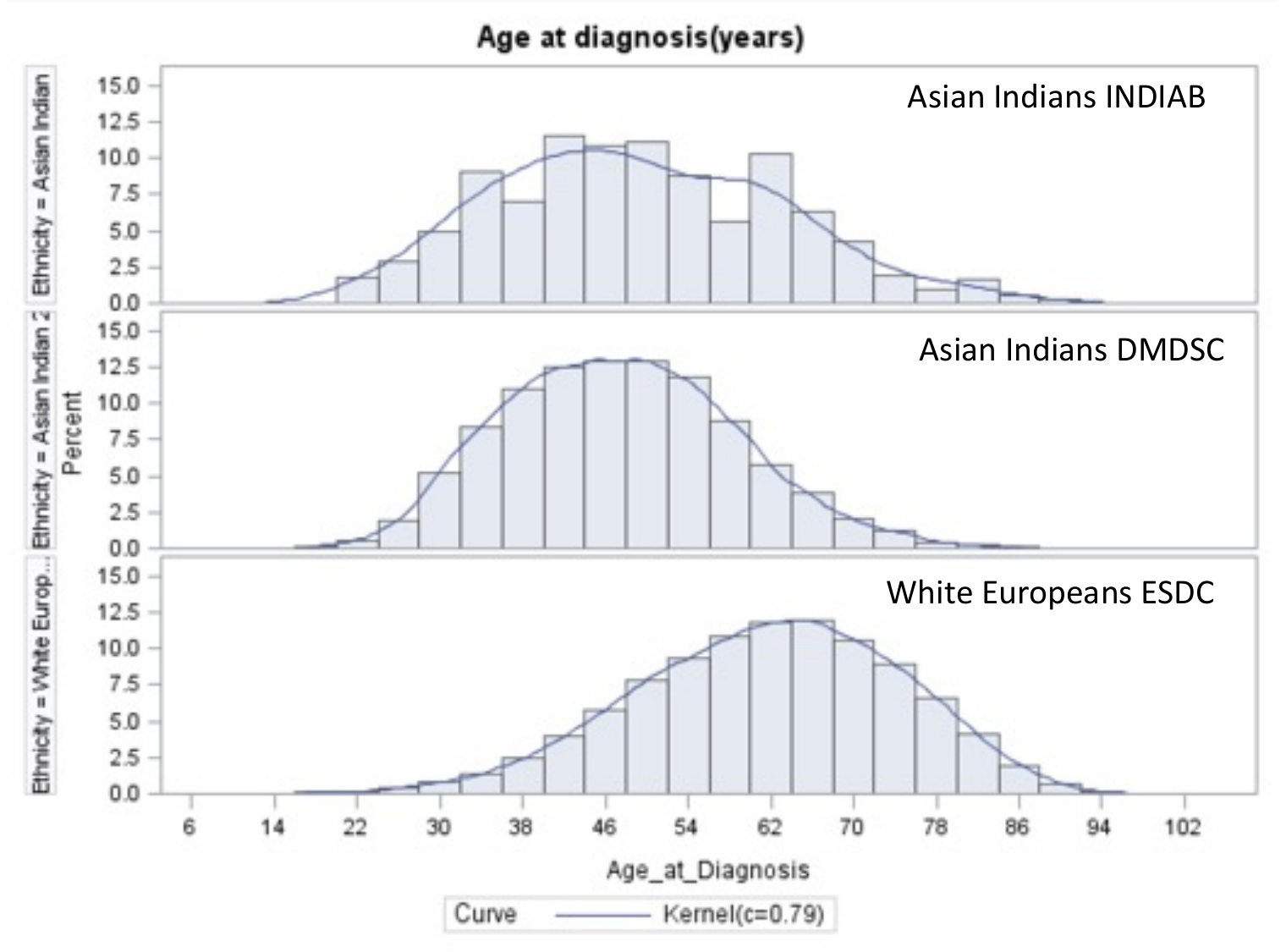

Supplementary Figure 1. Histogram of age of diabetes onset in Asian Indians from population sample - INDIAB (top panel), Asian Indians from a diabetes specialty clinic - DMDSC (middle panel) and white Europeans - ESDC (bottom panel). Median age of diagnosis for Asian Indians in INDIAB was 48 years, and in DMDSC was 47 years, while for white Europeans was 62 years.

Supplementary Table 1. Demographic characteristics of populations described in histograms

| **Trait\Population** | **White Europeans (ESDC) (n= 42,563)** | | **Asian Indian (INDIAB) (n = 1712)** | | **Asian Indians (DMDSC) (n=54,989)** | |
| --- | --- | --- | --- | --- | --- | --- |
|  | *Total n* | *Mean (SD)* | *Total n* | *Mean (SD)* | *Total n* | *Mean (SD)* |
| **Sex (Female)%** | 42,563 | 45% | 1,712 | 53% | 54,989 | 37.6% |
| **Average age (mean, SD) (years)** | 42,563 | 61.7 (13) | 1,712 | 50 (13) | 54,989 | 47.4 (11.2) |
| **Median age (median, IQR) (years)** | 42,563 | 62 (18) | 1,712 | 48 (21) | 54,989 | 47 (16) |
| **BMI (mean, SD) (Kg/m^2^)** | 39,234 | 32.3 (6.6) | 1712 | 24.2 (4.8) | 51,840 | 26.7 (4.5) |
| **Waist circumference (cm) (mean, SD)** | 8,046 | 106.1 (14) | 1681 | 86.2 (13) | 39,903 | 93.3 (10.6) |

ESDC: East Scotland Diabetes Cohort, INDIAB: Indian-Diabetes cohort, DMDSC: Dr. Mohan’s Diabetes Specialty Clinic

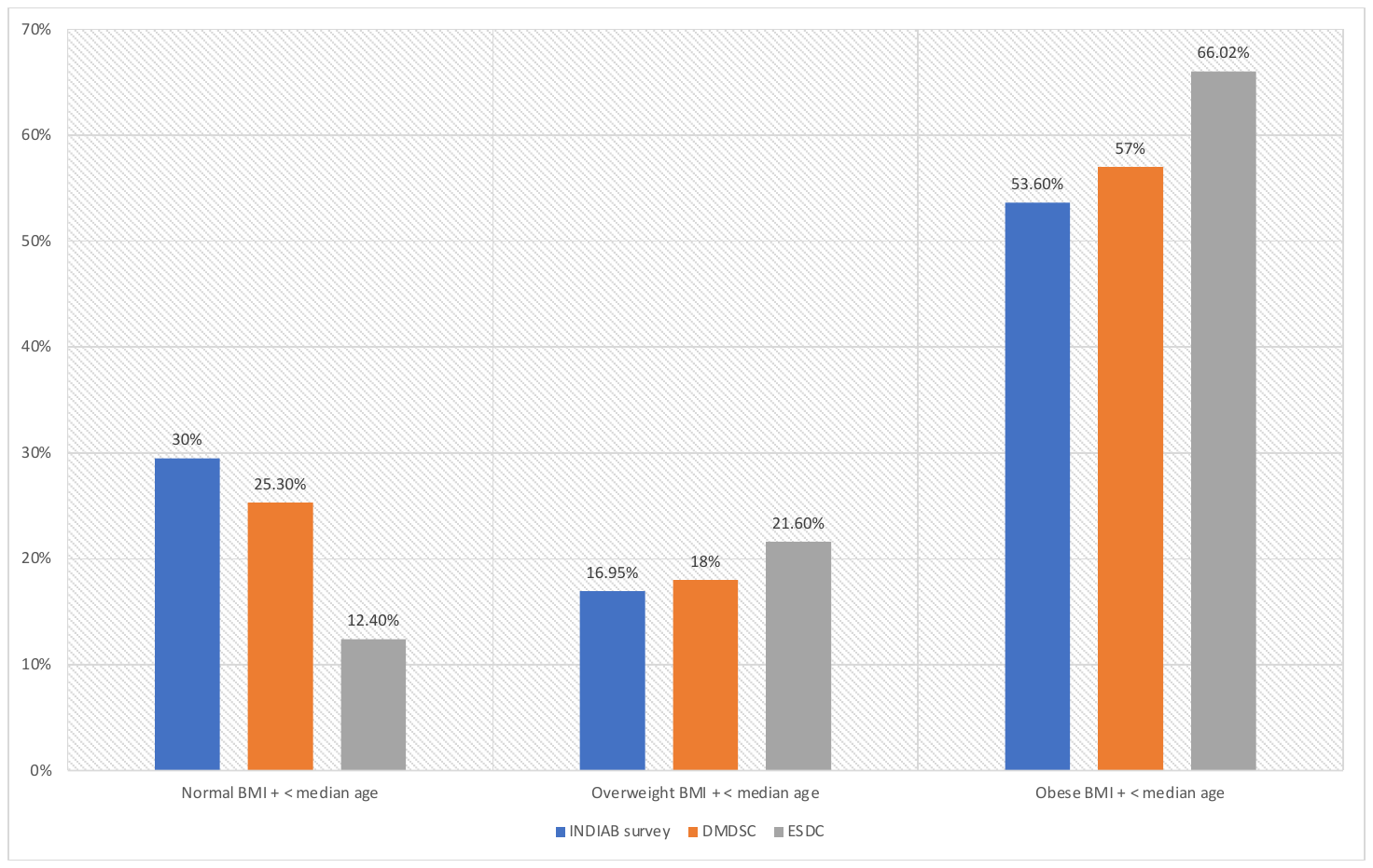

*Supplementary Figure 2. Sensitivity analyses using population-derived age cut-offs. Bar graphs of Asian Indians (blue -INDIAB and orange-DMDSC) and white Europeans (grey-ESDC) with early onset diabetes by proportion belonging to each BMI category. Normal BMI for Asian Indians was <23 kg/m2, overweight: 23-25kg/m2, obese: >25kg/m2. Normal BMI for white European population is <25 kg/m2, overweight 25-30 kg/m2 and obese >30 kg/m2 [16]. Since INDIAB and ESDC are a representative population, early or young onset diabetes in Asian Indians could also be defined as being under the age of 50 years, and under 62 for white Europeans.*

Supplementary Table 2. Examining trends across BMI categories in young (≤40 ) and old (>40) diagnosed Asian Indians

|  | **Young diagnosed (≤40 years)** | | | | **Older diagnosed (>40 years)** | | | |
| --- | --- | --- | --- | --- | --- | --- | --- | --- |
| **Variables** | **Normal BMI** | **Overweight BMI** | **Obese BMI** | **Test for trend,**  **P value** | **Normal BMI** | **Overweight BMI** | **Obese BMI** | **Test for**  **Trend,**  **P value** |
| **Numbers** | 3716 | 2723 | 8824 |  | 9395 | 7012 | 23319 |  |
| **Age at diagnosis (years) (mean, SD)** | 33.7 (4.5) | 34.1 (4.3) | 34.2 (4.3) | -2.03, 0.04 | 53.7 (9.4) | 52.8 (8.7) | 52.0 (8.1) | 3.8, 0.0002 |
| **Sex (% female)** | 24% | 25% | 33% | 10.9, <0.0001 | 33% | 33% | 46% | 24.3, <0.0001 |
| **BMI (kg/m^2^)  (mean, SD)** | 21.2 (1.6) | 24.1 (0.6) | 29.3 (3.7) | -- | 21.1 (1.7) | 24.1 (0.60) | 29.2 (3.6) | -- |
| **Waist circumference (females) (cm) (mean, SD)** | 77.23 (7.7) | 83.6 (6.5) | 94.0 (9.6) | 33.6, <0.0001 | 78.8 (7.4) | 84.8 (6.4) | 94.4 (9.3) | 63.2, <0.0001 |
| **Waist circumference (males) (cm) (mean, SD)** | 81.9 (6.3) | 88.3 (5.0) | 98.9 (9.1) | 67.6, <0.0001 | 84.0 (6.6) | 90.6 (5.2) | 100.7 (8.7) | 98, <0.0001 |
| **HbA1c (%) (mean, SD)** | 9.7 (2.7) | 9.3 (2.4) | 8.9 (2.1) | 5.6, <0.0001 | 9.12 (2.6) | 8.84 (2.3) | 8.56 (2.1) | 3.6, 0.0004 |
| **HbA1c (mmol/mol) (mean, SD)** | 83 (30) | 78 (26) | 73.2 (24) | -- | 76 (29) | 73 (25) | 70 (23) | -- |
| **Fasted c-peptide (pmol/mL) (median, IQR)** | 700 (400) | 900 (400) | 1100 (500) | -9.7, <0.0001 | 800 (400) | 1000 (500) | 1100 (500) | -6.5, <0.0001 |
| **Stimulated c-peptide (pmol/mL) (median, IQR)** | 1600 (1200) | 2000 (1400) | 2500 (1600) | -5.4, <0.0001 | 2000 (1400) | 2400 (1500) | 2800 (1600) | -6.1, <0.0001 |
| **Stimulated c-peptide adjusted for insulin sensitivity** | 417 (333) | 556 (392) | 732 (481) | 22.5, <0.0001 | 538 (366) | 675 (469) | 811 (473) | 23.1, <0.0001 |
| **HOMA-S (median, IQR)** | 48.5 (27.8) | 38.2 (20.7) | 31.3 (15.0) | 9.6, <0.0001 | 43.9 (25.2) | 37.8 (19.1) | 32.6 (15.6) | 7.4, <0.0001 |
| **HOMA-B (median, IQR)** | 31 (41.7) | 41.8 (44.2) | 56.8 (61.3) | -4.6, <0.0001 | 50.2 (64) | 61.4 (64.7) | 76.1 (72.1) | -4.0, <0.0001 |
| **Family history of diabetes (%)** | 63% | 67% | 73% | -11.9, <0.0001 | 36.9% | 43.0% | 47.6% | -17.4, <0.0001 |
| **LDL (mmol/L) (mean, SD)** | 2.97 (0.94) | 2.98 (0.90) | 2.99 (0.90) | -1.6, NS | 3.06 (0.96) | 3.04 (0.94) | 3.03 (0.92) | 1.3, NS |
| **HDL (mmol/L) (mean, SD)** | 1.03 (0.24) | 1.00 (0.21) | 0.98 (0.21) | -8.9, <0.0001 | 1.07 (0.26) | 1.05 (0.24) | 1.06 (0.24) | 0.92, NS |
| **Triglycerides (mmol/L) (median, IQR)** | 1.63 (1.30) | 1.81 (1.39) | 1.81 (1.33) | -4.9, <0.0001 | 1.57 (1.06) | 1.67 (1.08) | 1.63 (1.03) | -5.01, <0.0001 |
| **Total Cholesterol (mmol/L) (mean, SD)** | 4.94 (1.14) | 5.00 (1.21) | 4.94 (1.11) | -0.02, NS | 5.01 (1.2) | 4.99 (1.15) | 4.96 (1.11) | -3.6, 0.0001 |
| **ALT (IU/L) (median, IQR)** | 26 (21) | 31 (25) | 34 (29) | -6.2, <0.0001 | 22 (14) | 24 (16) | 25 (17) | NS |

Test for trend for parametric variables using linear regression (t statistic), non-parametric using Jonckheere-Terpstra test (Z statistic) and for categorical variables using Cochran-Armitage Trend test (Z statistic). Threshold for significance after correction for multiple testing = 0.001.

### Trends across BMI categories in young Asian Indians with diabetes

Young lean Asian Indians had earlier onset of diabetes, were more likely to be male, had higher HbA1c, lower fasted and stimulated C-peptides, lower beta cell function, and higher insulin sensitivity compared to those diagnosed young and with obese BMI. Using Asian India-specific and sex-specific waist circumference thresholds (<80 cm for females and <90 for males) (7), a higher proportion of males (94%) had normal waist circumference in the lean young group compared to females (82%). On average, young, lean individuals had lower triglycerides and slightly higher HDL-C compared to those who were overweight or obese suggesting that diabetes in the young, low BMI group was not selectively associated with dyslipidemia.

HOMA-B, a surrogate estimate of beta cell function, was 34.7% in young and lean Asian Indians and 46.4% and 64.1% in overweight and obese individuals respectively (Z statistic = 20, P value <0.0001). Similar results were seen for fasting and stimulated c-peptide. This suggests young and lean Asian Indians with new onset diabetes have lower beta cell function compared to those who are overweight or obese. HOMA-S a surrogate estimate of insulin sensitivity was higher (47.8% of health insulin sensitivity) in lean individuals compared to those who were overweight (38.3%) and obese (32%). Intriguingly, family history of diabetes in first degree relatives, an important risk factor of diabetes was most frequent in those diagnosed with higher BMI (65%) compared to those diagnosed with normal BMI (55%). This surprising trend was also seen in older diagnosed individuals.

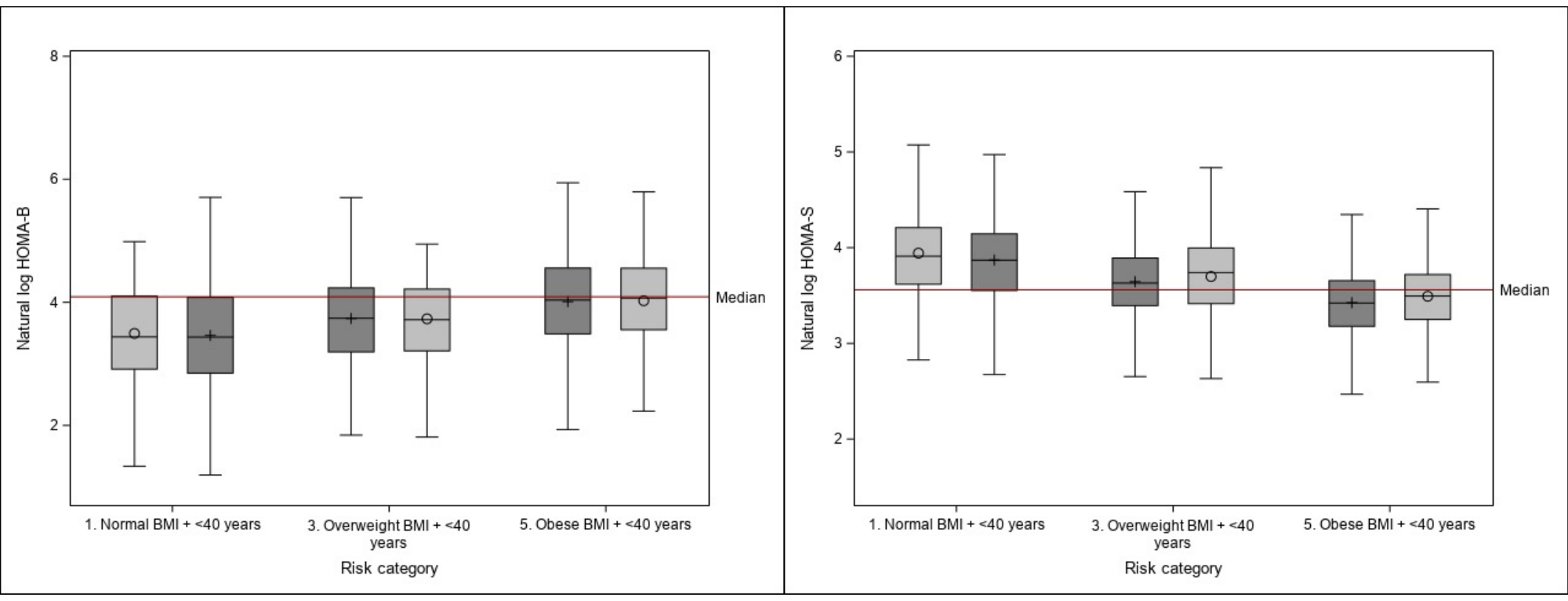

*Supplementary Figure 3. Differences in HOMA-B (left) and HOMA-S (right) (both natural log transformed) in those diagnosed young across BMI categories. Males in light gray, females in dark gray. After adjusting for insulin sensitivity (HOMA-S) the association between BMI categories and HOMA-B remained significant (P<0.0001).*

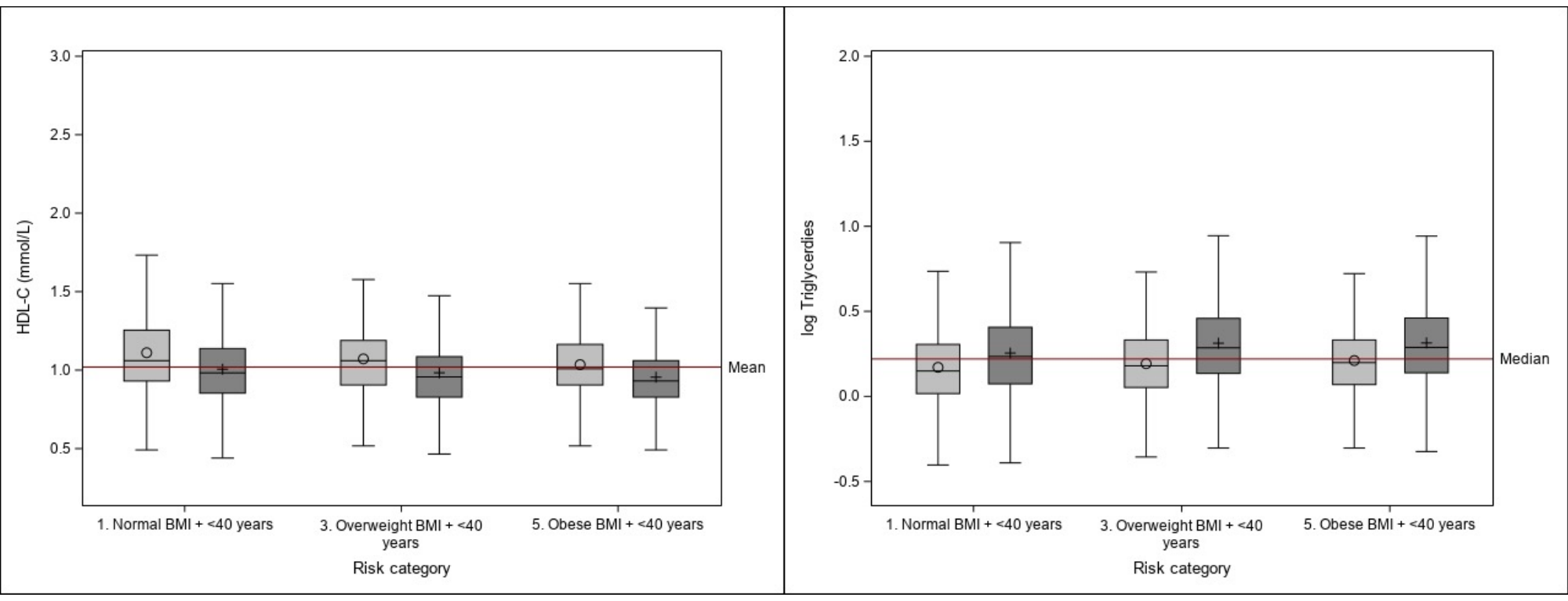

*Supplementary Figure 4. Differences in HDL-C (left) and triglycerides (log transformed) in those diagnosed young across BMI categories. Males in light gray, females in dark gray.*

Supplementary Table 3. Quantile regression models for glycemic traits showing adjusted differences across BMI groups in young diagnosed Asian Indians (diagnosis at age of 40 years or younger)

|  | **Median (and inter-quartile range) difference between successive BMI groups** | **Median (and inter-quartile range) difference between normal and obese groups** | **Test for trend across BMI categories** |
| --- | --- | --- | --- |
| **Fasting C-peptide levels (pmol/mL)** | 167 (131, 202) | 393 (362,424) | 9.2, <0.0001 |
| **Stimulated C-peptide levels (pmol/mL)** | 371 (240, 503) | 864 (763, 976) | 5.5, <0.0001 |
| **Stimulated C-peptide levels (pmol/mL) * adjusted for HOMA-S** | 208 (77, 340) | 492 (353, 616) | 3.1, 0.002 |
| **Differences in HOMA-S** | 10.2 (7.6, 12.7) | 16.97 (14.94, 18.66) | 7.8, <0.0001 |
| **Differences in HOMA-B** | 9.9 (5.17, 14.7) | 24.7 (20.2, 28.6) | 4.08, <0.0001 |
| **Differences in HOMA-B *adjusted for HOMA-S** | 9.9 (5.12, 14.8) | 25.16 (20.6, 28.6) | 4.04, <0.0001 |

Differences adjusted for sex, family history of type 2 diabetes, HDL-c levels. *Differences in HOMA-B and stimulated C-peptides were additionally adjusted for insulin sensitivity (HOMA-S). P value threshold after correction for multiple testing was 0.008.

Supplementary Table 4. Allele frequencies from variants in the insulin secretion/beta-cell function pPS in INSPIRED (ESDC and DMDSC) as well as UK Biobank cohorts.

| **Gene** | **SNP** | **Chromosome** | **Risk Allele** | **RAF INSPIRED-DMDSC** | **RAF INSPIRED-ESDC** | **RAF UKB South Asians** | **RAF UKB white Europeans** | **Susceptibility Beta HOMA-B** |
| --- | --- | --- | --- | --- | --- | --- | --- | --- |
| *MTNR1B* | rs10830963 | 11 | G | 0.34 | 0.28 | 0.32 | 0.25 | -0.039 |
| *GCK* | rs730497 | 7 | A | 0.11 | 0.19 | 0.19 | 0.17 | -0.025 |
| *TCF7L2* | rs7903146 | 10 | T | 0.30 | 0.32 | 0.29 | 0.3 | -0.02 |
| *ADCY5* | rs11708067 | 3 | A | 0.48 | 0.47 | 0.48 | 0.47 | -0.016 |
| *SLC30A8* | rs3802177 | 8 | C | 0.48 | 0.46 | 0.47 | 0.46 | -0.016 |
| *HNF1A* | rs1169288 | 12 | C | 0.34 | 0.31 | 0.31 | 0.27 | -0.0056 |
| *TMEM258* | rs102275 | 11 | T | 0.49 | 0.44 | 0.48 | 0.44 | -0.0054 |
| *ABO* | rs505922 | 9 | C | 0.3 | 0.29 | 0.35 | 0.27 | -0.0017 |

RAF: Risk allele frequency, INSPIRED-DMDMSC: the Asian Indian DMDMSC cohort, INSPIRED-ESDC: white European ESDC cohort, UKB refers to UKBiobank, susceptibility to HOMA-B calculated from estimates provided by Dupuis et al. (8)

Risk allel frequency

|  | INSPIRED Asian Indians | INSPIRED White Europeans | UK Biobank South Asian | UK Biobank White Europeans |
| --- | --- | --- | --- | --- |
| MTNR1B | 0.34 | 0.28 | 0.32 | 0.25 |
| GCK | 0.11 | 0.19 | 0.19 | 0.17 |
| TCF7L2 | 0.295 | 0.32 | 0.29 | 0.3 |
| ADCY5 | 0.48 | 0.47 | 0.48 | 0.47 |
| SLC30A8 | 0.476 | 0.46 | 0.47 | 0.46 |
| HNF1A | 0.34 | 0.31 | 0.31 | 0.27 |
| TMEM258 | 0.49 | 0.44 | 0.48 | 0.44 |
| ABO | 0.3 | 0.29 | 0.35 | 0.27 |

Supplementary Figure 5. Comparative risk allele frequencies (RAF) of insulin secretion/beta-cell function variants in the INSPIRED Asian Indian cohort (DMDSC – in blue) INSPIRED white European cohort (ESDC orange), UK Biobank South Asians (grey) and UK Biobank white Europeans (yellow). Variants are sorted in descending order from left to right based on effect size of susceptibility to poor beta cell function.

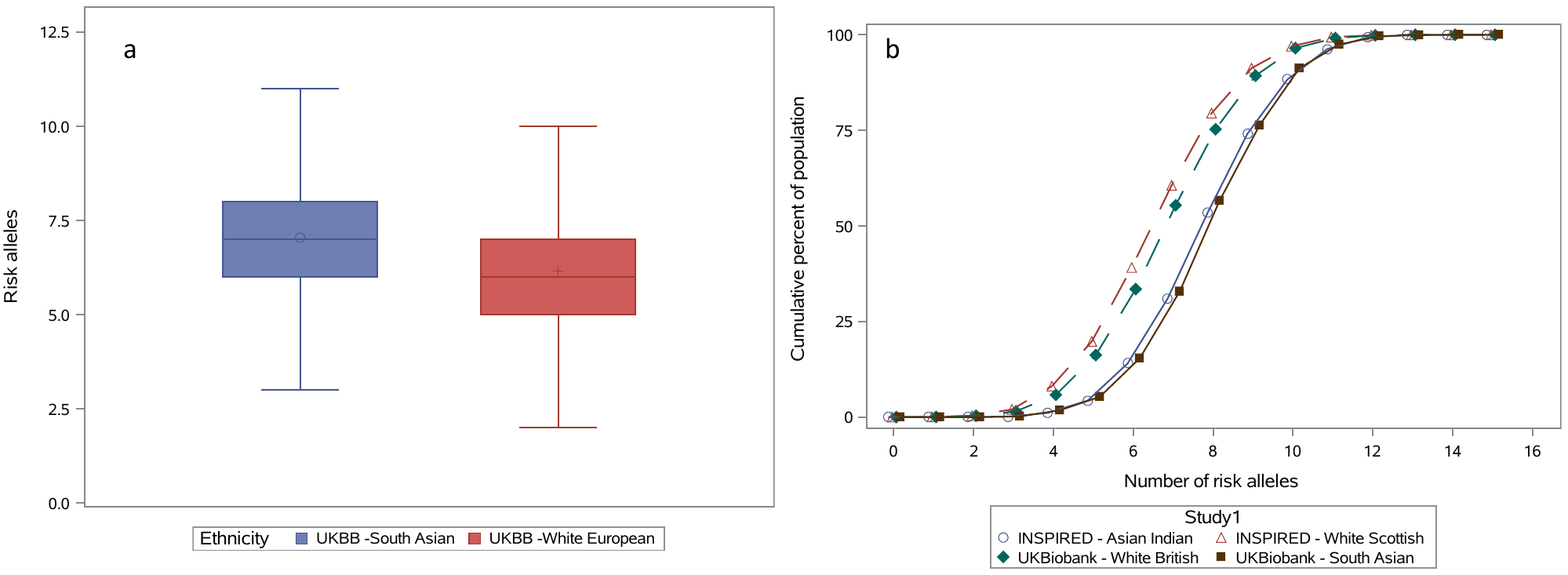

Supplementary Figure 6. Replication of difference in risk alleles for insulin secretion pPS in UKBiobank. Boxplots on the left (Fig 6a) show the distribution of risk alleles of insulin secretion of pPS in White Europeans (n = 17,183) and South Asians (n=1153) with type 2 diabetes in the UK Biobank. South Asians had significantly more risk alleles (Wilcoxon-Mann-Whitney P value < 0.0001). The empirical distribution of unweighted pPS also showed a significant difference using the Kolmogorov-Smirnov asymptotic two-sample test (P value<0.0001). Figure 6b the cumulative distribution of risk alleles in all studies is shown. INSPIRED samples are shown in open shapes (Asian Indians in blue circles and white Europeans from Scotland in red triangles) and UKBiobank in filled shapes (South Asians in brown squares and white Europeans from United Kingdom in green diamond). In the UK Biobank, the south Asian cohort (n=1153) is comprised predominantly of individuals of Asian Indian descent (n=700). The white population in UKBiobank is a mixture of individuals of English, Welsh, Irish, Scottish as well as other European descent living in England. In contrast the white Scottish population in ESDC are from the local population of Tayside, Scotland.

Supplementary Table 5. Test for normality of distributions for the beta-cell function weighted partitioned polygenic score

| **Measures of tendencies** | **White European pPS** | **Asian Indian pPS** |
| --- | --- | --- |
| **N** | 6933 | 5806 |
| **Skewness** | -0.38 | -0.13 |
| **Kurtosis** | -0.19 | -0.38 |
| **Mean (SD)** | -0.062 (0.035) | -0.102 (0.036) |
| **Median (IQR)** | -0.059 (0.048) | -0.102 (0.048) |
| **Mode** | -0.016 | -0.103 |

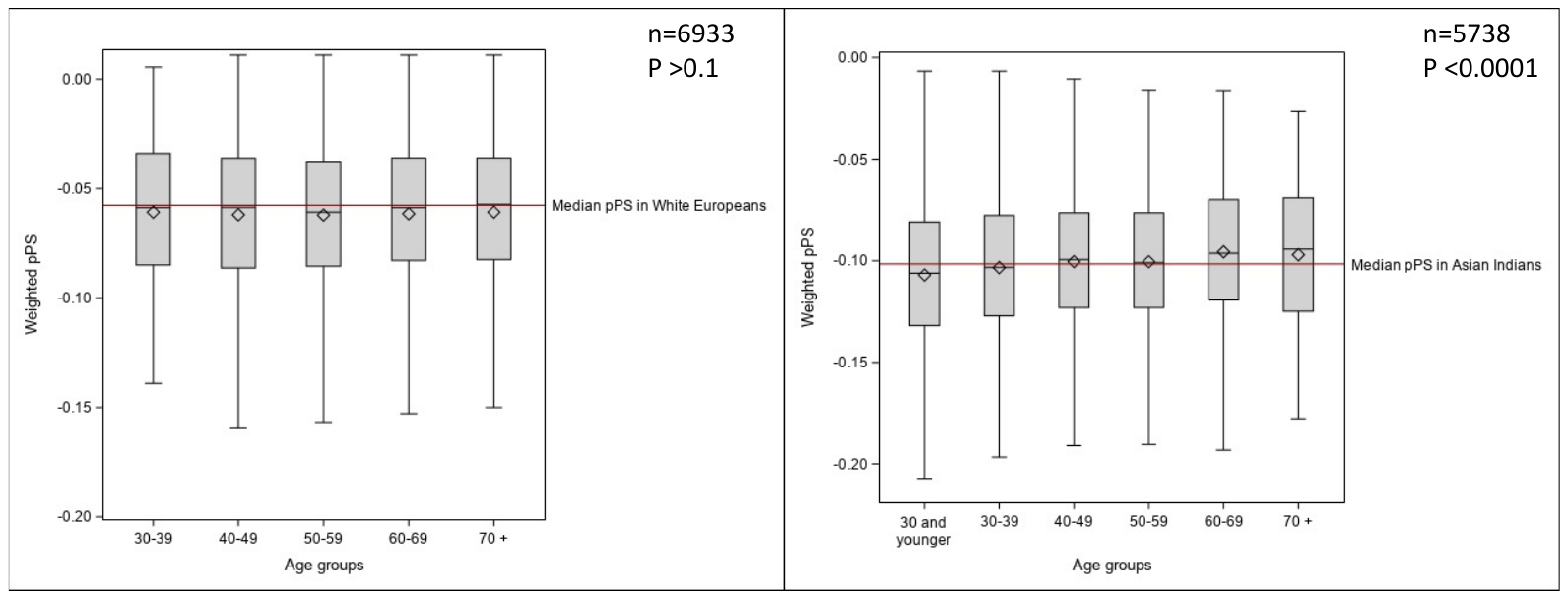

*Supplementary Figure 7. Boxplots demonstrating the association between weighted beta cell function/insulin secretion pPs and age of diagnosis in white Europeans (left) and Asian Indians (right). Using Jonckheere-Terpstra test for trend, the Z statistics for white Europeans was non-significant, while for Asian Indians was 5.3, P value <0.0001. There were insufficient numbers of white Europeans diagnosed below the age of 30 years.*

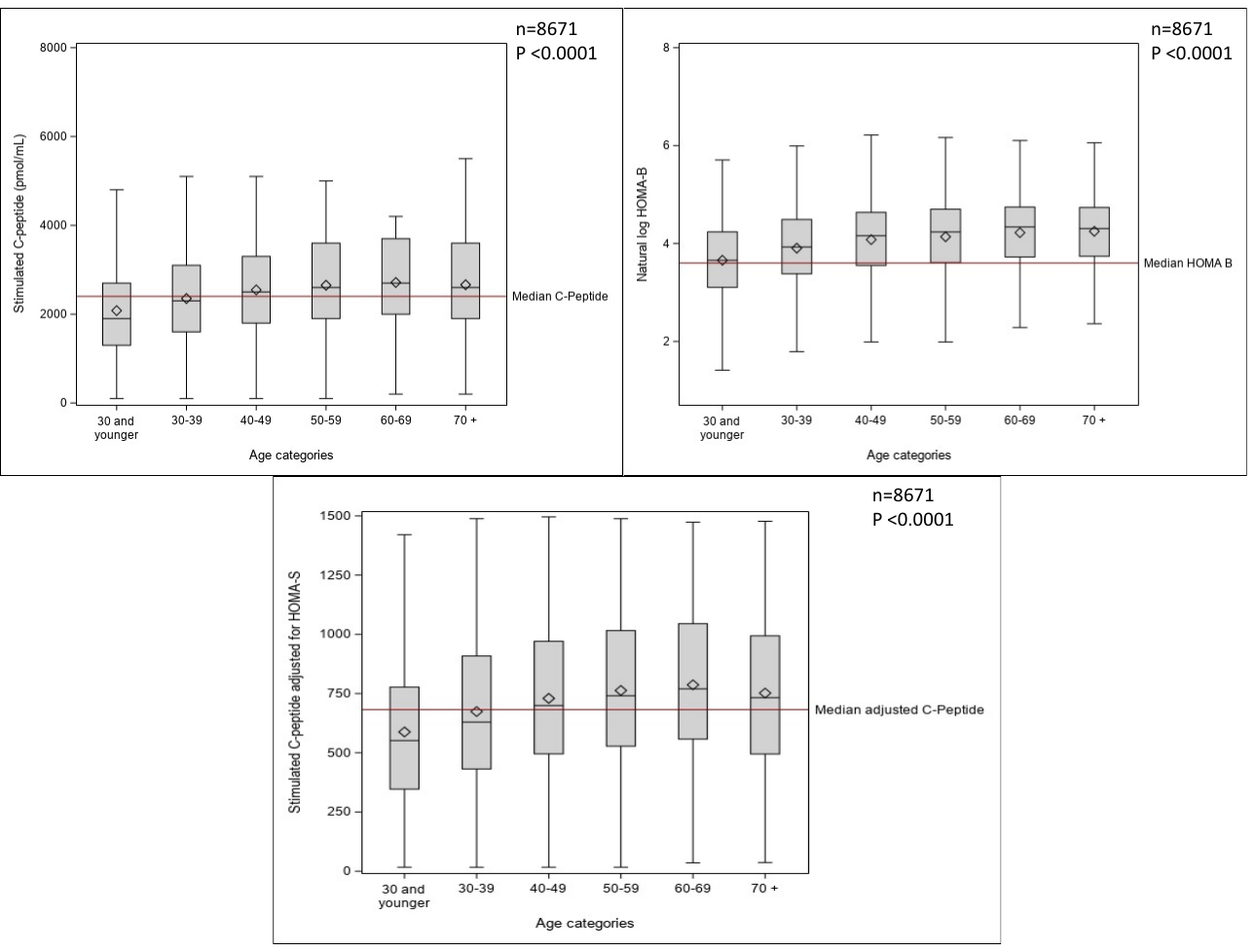

*Supplementary Figure 8. Trend of stimulated C-peptide, HOMA-B and HOMA-B adjusted for insulin sensitivity with age groups at diagnosis in Asian Indians. Using Jonckheere-Terpstra test for trend, the Z statistics for stimulated C-peptides was 17.1 and P value < 0.0001, for HOMA B was 18.99 and P value < 0.0001 and for stimulated c-peptides adjusted for insulin sensitivity was 15.7 and P value <0.0001.*

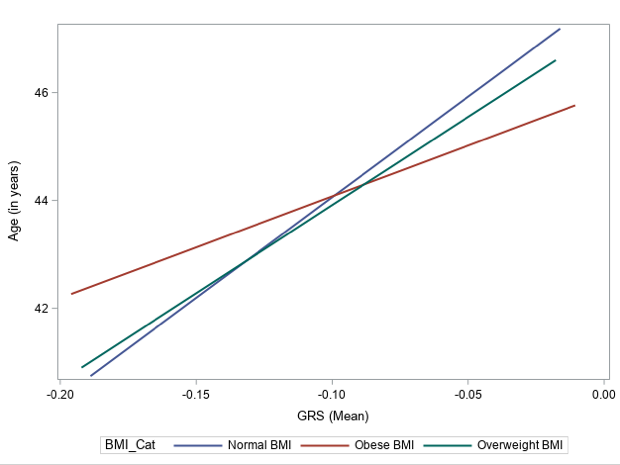

pPS

Supplementary Figure 9. Beta cell function partitioned polygenic risk score for beta cell function is a stronger predictor of age at diagnosis for those with normal BMI (in blue) compared to overweight (green) or obese BMI (red). Statistics provided in Supplementary Table 6.

Supplementary Table 6. Linear association of beta cell pPS and age at diagnosis of diabetes in Asian Indians stratified by BMI category.

| **Sub-groups** | **n** | **Beta for HOMA-B weighted GRS (SE)** | **P value** | **R^2^** |
| --- | --- | --- | --- | --- |
| **Normal BMI** | 409 | 37.31 (16.7) | 0.02 | 1.2 % |
| **Overweight BMI** | 257 | 32.68 (20.7) | 0.11 | 0.97% |
| **Obese BMI** | 847 | 18.9 (10.76) | 0.08 | 0.36% |
